# Validation of a New AI Seizure Detection Algorithm for Pre-term Neonates, Term Neonates and Infants

**DOI:** 10.1101/2025.10.31.25339264

**Authors:** Archit Gupta, Leonardo Tozzi, Tanaya Puranik, Suma Anand, Courtney Juliano, Maite La Vega-Talbott, Rana R. Said, Avantika Singh, Joseph E. Sullivan, Nicholas S. Abend, Cecil D. Hahn, Baharan Kamousi

## Abstract

Seizures are frequent in the first year of life and they are associated with increased risk of long-term neurological disability. Accurate diagnosis of seizures in neonates and infants requires continuous electroencephalogram (EEG) monitoring with expert interpretation. While rapid detection of seizures allows timely management and may influence neurodevelopmental outcomes, it may not be feasible in settings without round-the-clock expert interpretation.

We evaluated the real-world performance of Clarity (Ceribell Inc.), a novel algorithm for automated seizure detection from EEG of preterm neonates, term neonates, and infants in a validation dataset of patients of postnatal age 0-1 years and gestational age 22-42 weeks who underwent continuous EEG monitoring at three hospitals in the United States. We quantified algorithm performance using the following metrics: (1) area under the curve for classification of 10-second EEG segments as seizure or no seizure; (2) sensitivity and false positives in detecting seizure episodes; and (3) sensitivity, specificity and negative/positive predictive value in identifying recordings where seizure duration reached or exceeded predetermined thresholds.

The validation dataset consisted of 756 EEG recordings with a cumulative duration of 5230 hours (median: 2.94 hours, interquartile interval: 1.46-12.62 hours) from 167 preterm neonates, 323 term neonates, and 266 infants. A total of 568 seizures occurred in 75 recordings, with a cumulative seizure duration of 15.13 hours. The algorithm achieved an area under the curve of 0.96. The algorithm was correct 99-100% of the time when ruling out seizures in a recording. It correctly detected 75-83% of seizure episodes and identified 89-94% of recordings containing seizures. Performance was equivalent between preterm neonates, term neonates, and infants and across the three participating hospitals.

Overall, the Clarity algorithm demonstrated high performance in detecting and ruling out seizures from EEG data in preterm neonates, term neonates, and infants across three hospitals. This demonstrates real-world feasibility and high reliability of automatic seizure detection with Clarity for continuous EEG monitoring in the first year of life.

## Introduction

Seizures are common in the first year of life ^1^ and are especially frequent in the neonatal period, occurring in approximately 1.5–5.5 per 1,000 live births ^2^. Seizures in neonates are associated with mortality rates of up to 20% and confer an increased risk of long-term neurological disability, including epilepsy, cerebral palsy, and cognitive impairment ^2–9^. Importantly, a higher duration of seizure activity (or “seizure burden”) is associated with greater mortality and morbidity ^5,10–15^. Therefore, both rapid detection and early treatment of seizures are critical to minimize injury to the developing brain ^13,16,17^.

Electroencephalography (EEG) is the gold standard for diagnosing seizures ^18,19^. Use of EEG is particularly important in neonates since seizures in this population are often electrographic only without clinical correlates. Additionally, atypical movements are common and can be mistaken for seizures ^20–24^. Therefore, current guidelines recommend continuous EEG monitoring in neonatal intensive care units (NICUs) for patients at high risk for seizures ^23,25^. However, diagnosing seizures in neonates and infants using EEG requires highly specialized experts who are not available at all times in many NICUs. One recent multicenter study found that even with continuous EEG in place, only 11% of neonatal seizures were recognized and treated by clinicians within one hour of onset ^26^.

Recently, advances in machine learning and artificial intelligence have enabled the development of algorithms for automated seizure detection from the EEG signal in neonates and infants. Early algorithms relied on the summarization of EEG data into a set of hand-crafted features ^27–29,29–32^, whereas more recent models rely on neural networks and are able to operate directly on raw EEG data ^33–36^. These algorithms are promising tools to efficiently and rapidly identify and manage seizures. In a recent multicenter randomized trial, these approaches increased the total number of seizure hours detected in neonates ^37^. However, published algorithms have limitations that impact their deployment in clinical practice. First, validation datasets used in previous publications are mostly small (<100 patients) so generalizability is uncertain. Second, previous algorithms have not been trained and evaluated using channel-level labels generated by board-certified pediatric electroencephalographers. Given pediatric electroencephalographers undergo extensive training to accurately identify seizures from neonate and infant EEGs, their interpretations are the gold standard for generating labels to train an automated seizure detection algorithm. Third, public datasets that are most commonly used for model validation do not include preterm neonates (e.g. ^38,39^). Preterm neonates are heavily represented in the NICU and may have a higher risk of seizures ^2^. Also, seizures in preterm neonates can have a distinct EEG morphology compared to seizures in term neonates and infants ^40^. Therefore, it is critical that any algorithm be evaluated in preterm neonates.

We performed a study to assess the real-world performance of Neonate and Infant Clarity (hereafter “Clarity”, Ceribell Inc.), a novel automated seizure detection algorithm, in preterm neonates, term neonates, and infants. We leveraged a large multi-site validation dataset from three hospitals within the United States, annotated with channel-level labels provided by board-certified pediatric electroencephalographers. To assess generalizability across ages and clinical settings, we also evaluated performance separately for preterm neonates, term neonates, and infants as well as separately for each hospital.

## Methods

### Validation dataset

The validation dataset was obtained retrospectively and consisted of 756 continuous EEG monitoring recordings from 715 patients of postnatal age 0-1 years and gestational age 22-42 weeks, yielding 5,230 hours of EEG data. The data was sourced from three hospitals to test generalizability of the algorithm across a diverse population, including Children’s Wisconsin (Medical College of Wisconsin, Wisconsin), Children’s Medical Center Dallas (University of Texas Southwestern, Texas), and Kravis Children’s Hospital (Mount Sinai Hospital, New York).

Clarity uses as input EEG data from the 12 bipolar channels of the recommended neonatal montage defined by American Clinical Neurophysiology Society: Fp1-T3, T3-O1, Fp1-C3, C3-O1, Fp2-T4, T4-O2, Fp2-C4, C4-O2, T3-C3, C3-Cz, Cz-C4, C4-T4 ^41^. Any additional channels were discarded. The validation dataset did not contain any patient that was part of the dataset used to train Clarity.

### Dataset labeling

Expert review of EEG is the gold standard in identification of seizures in EEG waveforms ^42,43^. All EEGs were annotated by two or more board-certified pediatric electroencephalographers who were neurologists with fellowship training in epilepsy or clinical neurophysiology (see Supplementary Table 1 for qualifications and years of experience of the reviewers). When the first two reviewers disagreed, additional reviewers were used until a majority consensus was reached. The reviewers annotated seizures according to established criteria ^44^. Within each EEG recording, reviewers used a custom annotation tool to mark the start and end times of individual seizures or other abnormal rhythms, and to specify the channels involved. The reviewers assessed a randomized assortment of EEGs across different sites to avoid rater-site interaction bias. The reviewers were not presented with the results of other reviewers or the result of Clarity, preventing them from being biased by either source. The reviewers completed EEG labeling before any validation testing was run on that data (i.e., prior to the generation of results from the algorithm). Therefore, all of the EEG reviews were blinded, and it was not possible for reviewers to bias their review in a way that affected the outcome of the validation testing. To mitigate any effects of inter-rater variability, the majority label from all reviewers was selected as the final label for each seizure event.

### Seizure detection algorithm

Clarity combines sequence models evaluating the evolution of each EEG channel in time with a convolutional neural network evaluating EEG patterns across channels. The algorithm classifies each 10-second EEG segment as either “seizure” or “no seizure”. These binary outputs are then continuously converted into a seizure burden for clinical use, defined as the percentage of time spent in seizure within the preceding 5-minute moving window. This seizure burden is updated every 10 seconds (Figure 1). Clarity generates an alert when the seizure burden is equal to or greater than a user defined threshold. Users can set this threshold based on the minimum seizure burden they deem clinically significant, allowing customization for different use cases.

**Figure 1:**
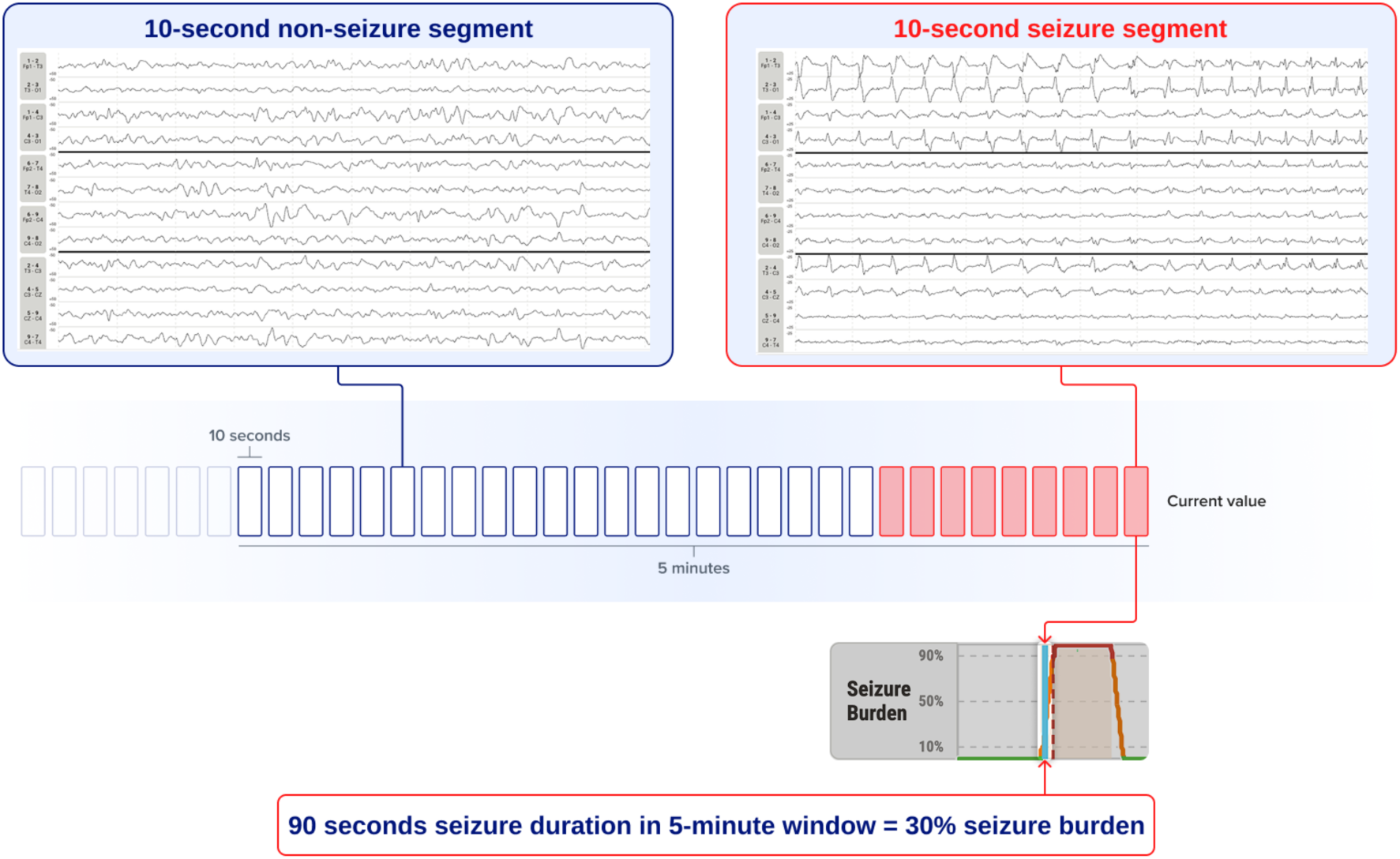
Output of the Clarity seizure detection algorithm. The algorithm classifies each 10-second EEG segment as seizure or no seizure. These binary outputs are then continuously converted to a seizure burden for clinical use, defined as percentage of seizure activity within the prior 5 minutes. Users can also set an alert when the seizure burden crosses a certain threshold, allowing customization for different use cases.

The algorithm was previously trained on a dataset of 402 EEGs from 338 patients collected in 8 hospitals and labeled for seizures as described above. The validation dataset presented here was collected separately and did not contain any patient that was part of the training dataset.

### Seizure detection algorithm evaluation

Consistent with current best-practice frameworks for the evaluation of seizure detection algorithms, we evaluated the performance of Clarity by conducting analyses at the segment level, episode level, and recording level ^45^.

In the segment level analysis, we quantified the ability of the algorithm to correctly classify these segments by calculating the area under the curve (AUC) for the binary classification of the 10 second EEG segments classified by Clarity as seizure or no seizure.

In the episode level analysis, we evaluated the algorithm’s ability to detect seizure episodes, defined as an interval in which the true seizure burden is elevated above a specified threshold. To define the start and end of these episodes, we used the true seizure burden calculated within a 5-minute window. When determining the maximum seizure burden of the episode, we considered the maximum seizure burden in a window starting 30 seconds before and ending 30 seconds after the true seizure episode. We also merged consecutive seizures with a gap of less than 120 seconds into a single seizure. This was done to account for slight misalignments in seizure annotation and prediction due to gradual changes in EEG at the beginning and end of seizures or to inter-rater variation in marking start and end of seizures. We evaluated the ability of the algorithm to detect seizures whose duration reached or exceeded three thresholds in any 5-minute window: ≥30 seconds, ≥150 seconds and ≥270 seconds (i.e., ≥10%, ≥50% and ≥90% seizure burden within a 5-minute window). We considered a seizure detected when the estimated seizure burden was equal or greater than the true seizure burden of the episode. For each threshold, we calculated the percentage of seizures detected (sensitivity) and the number of false positives per hour.

In the recording-level analysis, we evaluated the algorithm’s ability to correctly identify recordings where seizure duration reached or exceeded three thresholds in any 5-minute window: ≥30 seconds, ≥150 seconds and ≥270 seconds (i.e. ≥10%, ≥50% and ≥90% seizure burden). To do this, we extracted the true maximum seizure burden and the maximum seizure burden predicted by the algorithm from each EEG recording. We considered a seizure recording detected when the maximum estimated seizure burden was equal or greater than the true maximum seizure burden of the recording. We calculated sensitivity, specificity, and negative predictive value of the algorithm at each threshold. Then, we calculated a second set of algorithm performance metrics after excluding recordings that contained seizures with shorter maximum seizure duration than the predefined threshold. This was done to account for cases where the algorithm correctly detected a seizure but overestimated its duration. An alert in this case might still be clinically relevant and may not be considered a false positive. In this second set of metrics, for instance, given a ≥270 seconds threshold, a recording with 150 seconds maximum seizure duration with an algorithmically predicted maximum seizure duration of 270 seconds was not counted as a false positive but instead removed from the calculation.

We also evaluated algorithm performance separately in pre-term neonates (PMA <37 weeks), term neonates (PMA 37-44 weeks), and infants (PMA ≥44 weeks). It was important to distinguish these groups because the different stages of brain development after birth are reflected in EEG changes ^19^. In particular, seizures can be challenging to identify in pre-term neonates since they can have lower voltage and frequency compared to those in full-term neonates ^40^. To evaluate whether the algorithm performance was robust across the sites, we also repeated our analyses for each site separately.

## Results

### Reference standard

The validation dataset consisted of 756 EEG recordings from 715 patients, with a cumulative recording duration of 5,230 hours (median duration: 2.94 hours, interquartile interval: 1.46-12.62 hours). Based on the age of patients at the time of EEG, 167 recordings were from preterm neonates, 323 from term neonates, and 266 from infants. No patients were excluded. Expert consensus determined that 75/756 (10%) recordings contained seizures and that 71/715 (10%) of patients had at least one seizure. There was a total of 568 seizures, yielding a cumulative seizure duration of 15.13 hours. The median seizure duration was 60 seconds (interquartile interval: 30-120 seconds) (Table 1).

**Table 1:** Sample demographics and seizure information. Abbreviations: h=hours, IQR=inter-quartile interval, PMA=post-menstrual age, SD=standard deviation, s=seconds

|  | All neonates and infants | Pre-term neonates (PMA <37 weeks) | Term neonates (PMA 37-44 weeks) | Infants (PMA >44 weeks) | Children's Medical Center Dallas | Kravis Children's Hospital | Children's Wisconsin |
| --- | --- | --- | --- | --- | --- | --- | --- |
| Number of patients | 715 | 157 | 321 | 237 | 175 | 249 | 291 |
| Number of recordings | 756 | 167 | 323 | 266 | 175 | 249 | 332 |
| Gestational age weeks mean ± SD | 36.40 ± 3.95 | 31.97 ± 4.02 | 38.22 ± 1.62 | 36.98 ± 3.78 | 38.32 ± 1.97 | 36.33 ± 4.09 | 35.45 ± 4.25 |
| PMA mean weeks ± SD | 45.80 ± 13.51 | 33.18 ± 3.18 | 39.55 ± 1.69 | 61.32 ± 11.01 | 47.98 ± 13.04 | 44.20 ± 12.09 | 45.86 ± 14.61 |
| Sex |  |  |  |  |  |  |  |
| Female | 341 (45%) | 65 (39%) | 151 (47%) | 125 (47%) | 86 (49%) | 117 (47%) | 138 (42%) |
| Male | 404 (53%) | 91 (54%) | 172 (53%) | 141 (53%) | 89 (51%) | 132 (53%) | 183 (55%) |
| Unknown | 11 (1.4%) | 11 (7%) | 0 (0%) | 0 (0%) | 0 (0%) | 0 (0%) | 11 (3%) |
| Race |  |  |  |  |  |  |  |
| American Indian or Alaska native | 2 (<1%) | 1 (1%) | 1 (<1%) | 0 (0%) | 0 (0%) | 0 (0%) | 2 (1%) |
| Asian | 26 (3%) | 5 (3%) | 10 (3%) | 11 (4%) | 1 (1%) | 18 (7%) | 7 (2%) |
| Black or African American | 143 (19%) | 35 (21%) | 62 (19%) | 46 (17%) | 39 (22%) | 42 (17%) | 62 (19%) |
| White | 350 (46%) | 80 (48%) | 136 (42%) | 134 (50%) | 50 (29%) | 88 (35%) | 212 (64%) |
| Other | 175 (23%) | 14 (8%) | 92 (28%) | 69 (26%) | 85 (49%) | 71 (29%) | 19 (6%) |
| Unknown | 60 (8%) | 32 (19%) | 22 (7%) | 6 (2%) | 0 (0%) | 30 (12%) | 30 (9%) |
| Ethnicity |  |  |  |  |  |  |  |
| Hispanic or Latino | 196 (26%) | 24 (14%) | 89 (28%) | 83 (31%) | 74 (42%) | 52 (21%) | 70 (21%) |
| Not Hispanic or Latino | 464 (61%) | 104 (62%) | 195 (60%) | 165 (62%) | 98 (56%) | 132 (53%) | 234 (70%) |
| Unknown | 96 (13%) | 39 (23%) | 39 (12%) | 18 (7%) | 3 (2%) | 65 (26%) | 28 (8%) |
| Total recording duration, h | 5230 | 753 | 2747 | 1730 | 2475 | 779 | 1976 |
| Median recording duration, h (IQR) | 2.94 (1.46-12.62) | 2.18 (1.21-3.54) | 4.10 (1.72-14.30) | 2.82 (1.39-12.30) | 13.30 (12.10-18.50) | 1.76 (1.25-3.36) | 2.68 (1.15-6.65) |
| Recordings with seizures | 75 (10%) | 17 (10%) | 40 (12%) | 18 (7%) | 24 (14%) | 13 (5%) | 38 (11%) |
| Median seizure duration, s (IQR) | 60 (30-120) | 50 (20-90) | 70 (30-120) | 60 (40-120) | 60 (40-100) | 40 (20-115) | 70 (30-145) |
| Cumulative seizure duration, h | 15.13 | 3.19 | 8.03 | 3.91 | 4.55 | 1.99 | 8.59 |

### Segment-level performance

Figure 2 illustrates Clarity performance in a sample recording. When evaluating seizure detection performance at the 10-second EEG segment level, the algorithm achieved an AUC=0.96 (Table 2). Performance was equivalent across subgroups of preterm neonates (AUC=0.93), term neonates (AUC=0.97), and infants (AUC=0.97). Performance was also equivalent across the three sites (Children’s Medical Center Dallas AUC = 0.96, Kravis Children’s Hospital AUC = 0.96, Children’s Wisconsin AUC = 0.96, Table 2).

**Figure 2:**
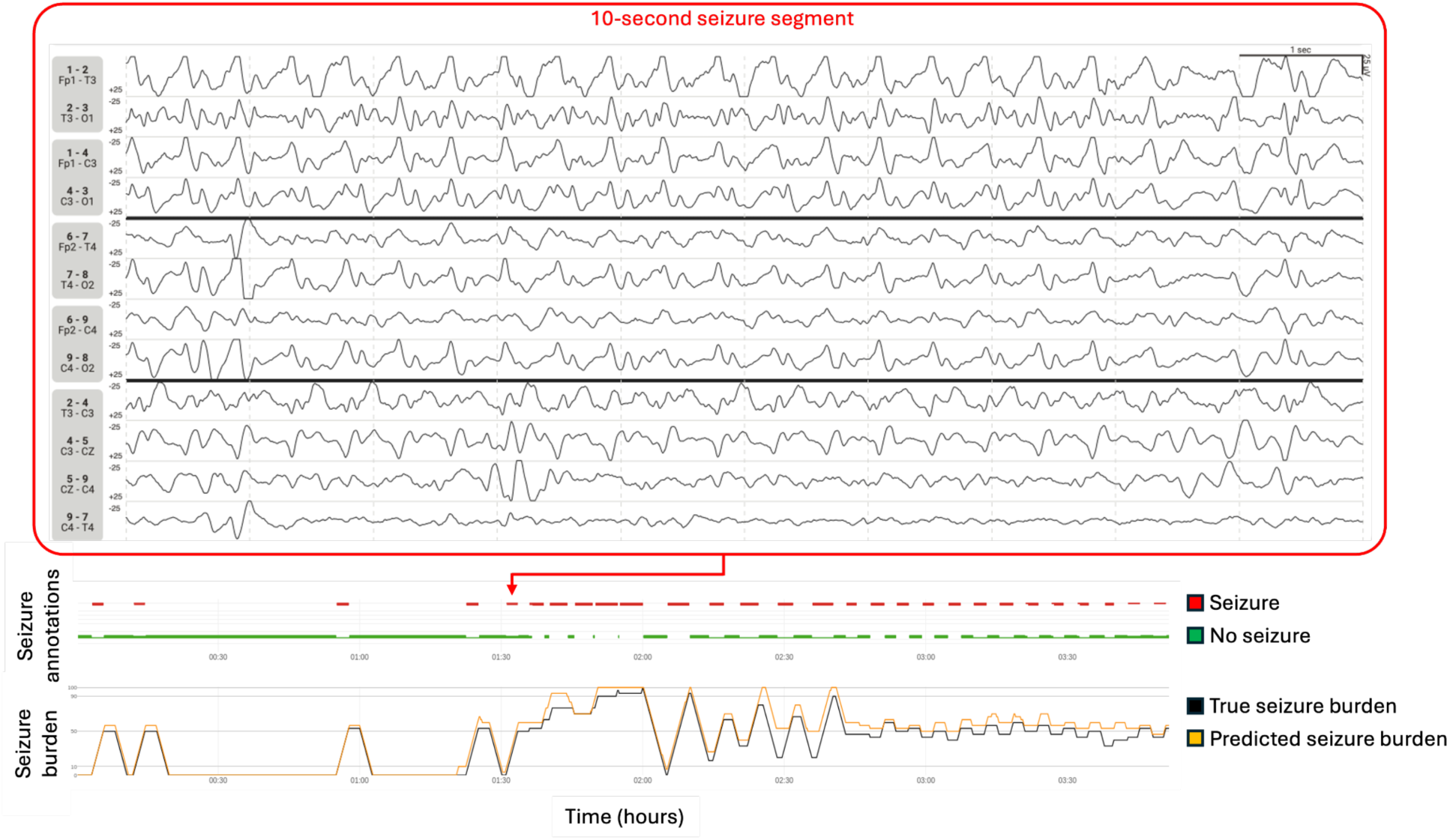
Seizure detection performance in a sample recording. Seizure annotations by electroencephalographers were used to calculate a ‘true’ seizure burden, defined as the percentage of time spent in seizure within the preceding 5-minutes. A 10-second EEG segment is depicted which had been annotated as containing a seizure, with a red arrow indicating its position on the summary of all seizure annotations. The calculated true seizure burden (black) and algorithmically predicted seizure burden (yellow) are illustrated below.

**Table 2:**
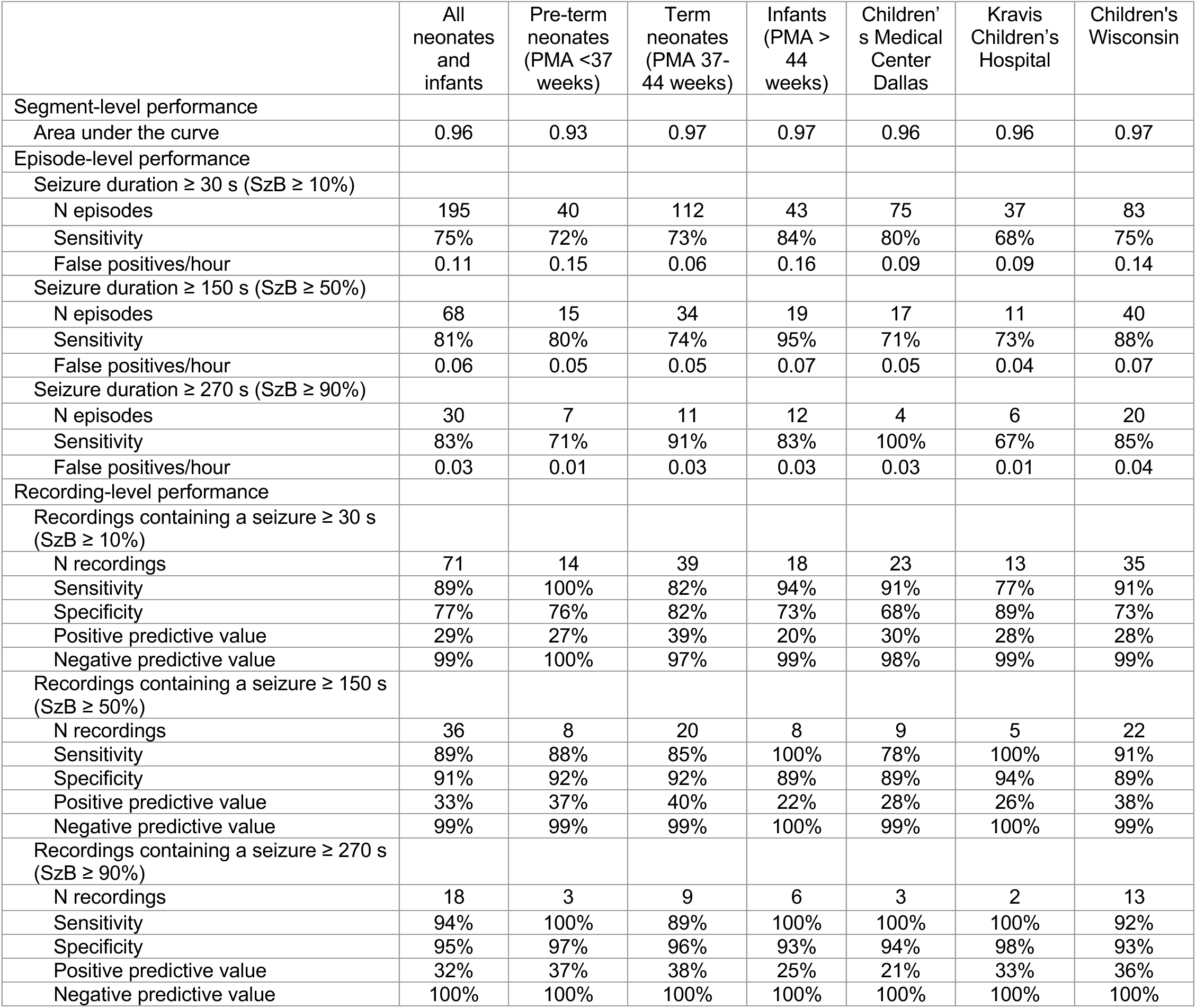
Algorithm performance. Clarity performance at varying levels of seizure duration in the whole sample and stratified by post-menstrual age and study site. Abbreviations: PMA=post-menstrual age, s=seconds, SzB=seizure burden.

### Episode-level performance

The algorithm correctly detected seizures with a duration ≥30 seconds in 75% of individual seizure episodes (sensitivity) while generating 0.11 false positives per hour (Table 2, Figure 3). For seizure durations of ≥150 seconds or ≥270 seconds, the algorithm sensitivity was consistently >80% and false positives per hour were consistently <0.10 (Table 2, Figure 3).

**Figure 3:**
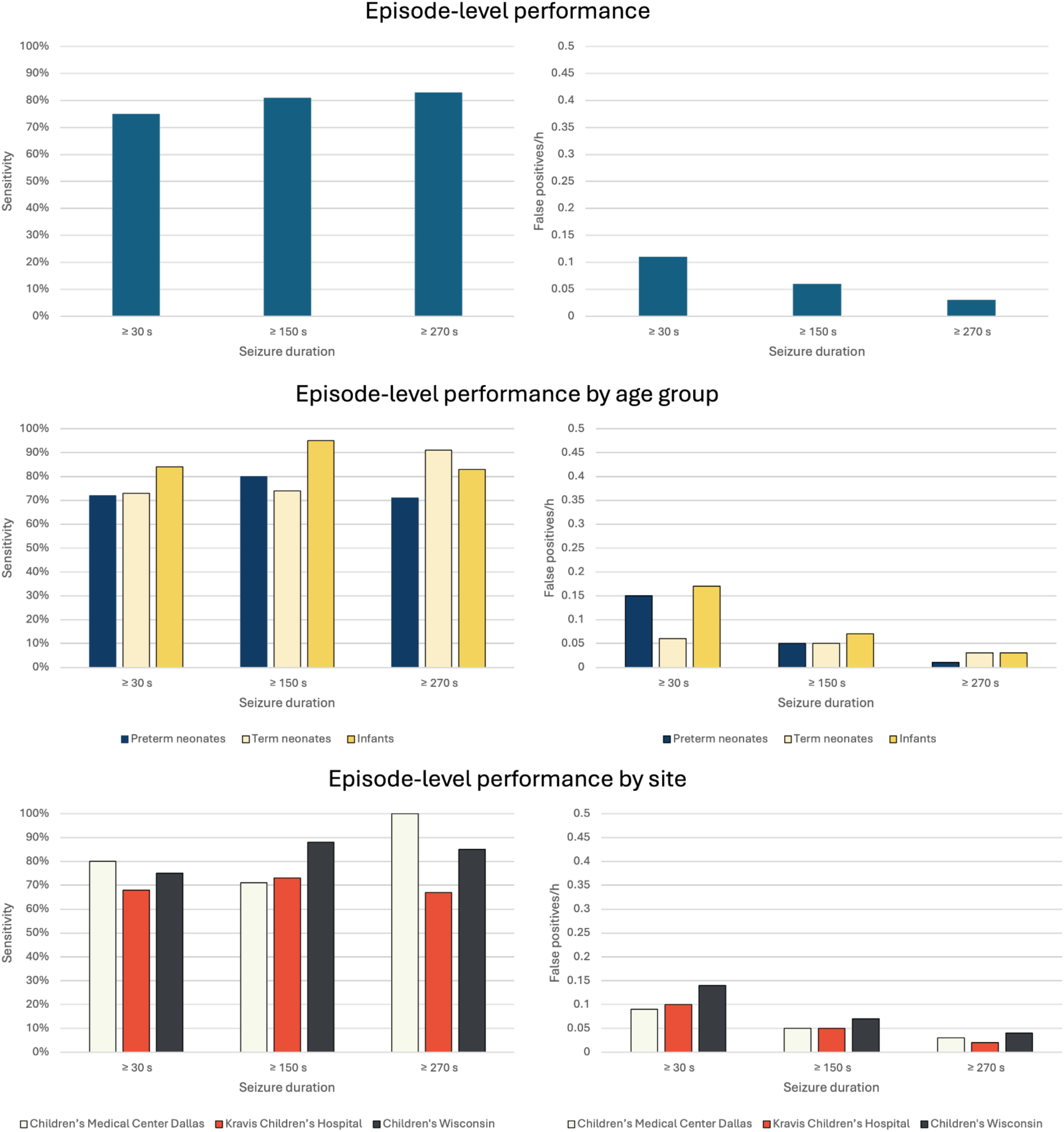
Episode-level algorithm performance. We evaluated the algorithm’s ability to detect seizure episodes with duration equal or greater than three thresholds: ≥30 seconds, ≥150 seconds and ≥270 seconds (i.e., ≥10%, ≥50% and ≥90% seizure burden in 5-minutes). For each threshold, we calculated sensitivity and false positives/hour. Data are presented for the whole sample and stratified by postmenstrual age and study site.

In preterm neonates, term neonates, and infants analyzed separately, sensitivity was 70%-95% depending on seizure duration. Performance was consistent when considering sites separately, with the exception of a slightly lower sensitivity at Kravis Children’s Hospital for a seizure duration of ≥30 seconds (68%) or ≥270 seconds (67%) (Table 2, Figure 3).

### Recording-level performance

For detecting recordings containing a seizure lasting ≥30 seconds in any 5-minute window, the algorithm sensitivity was 89%, specificity 77%, and negative predictive value 99%. For detecting recordings containing a seizure lasting ≥150 seconds, the algorithm sensitivity was 89%, specificity was 91%, and negative predictive value was 99% (Table 2, Figure 4). For detecting recordings containing a seizure lasting ≥270 seconds, the algorithm sensitivity was 94%, specificity was 95%, and negative predictive value was 100% (Table 2, Figure 4). When not considering as false positives cases where the algorithm correctly detected a seizure but overestimated its duration, specificity increased across all seizure burden thresholds (Supplementary Table 2).

**Figure 4:**
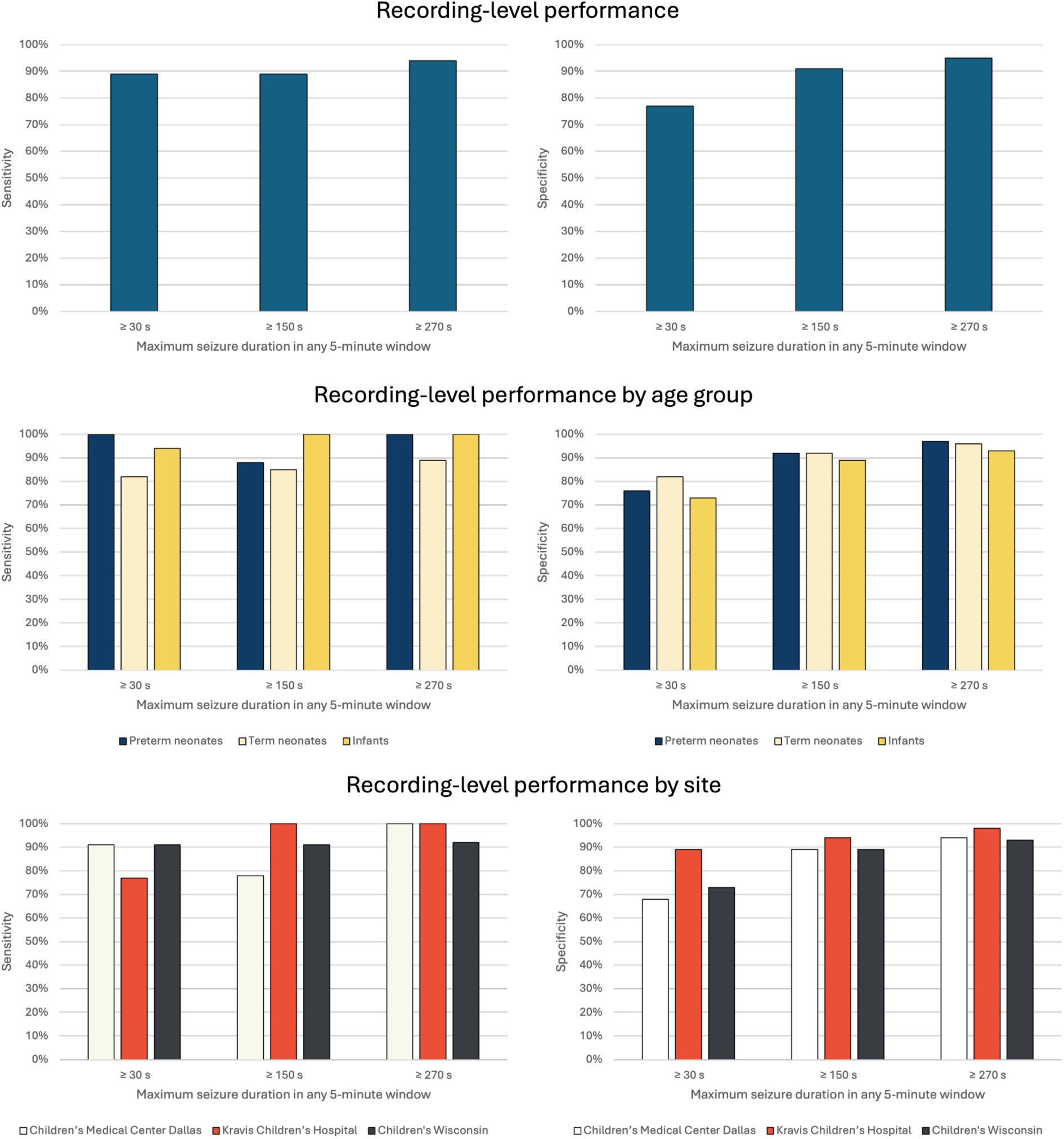
Recording-level algorithm performance. We evaluated the algorithm’s ability to identify recordings containing a seizure in any 5-minute window lasting ≥30 seconds, ≥150 seconds and ≥270 seconds (i.e., ≥10%, ≥50% and ≥90% seizure burden in 5-minutes). For each threshold, we calculated sensitivity and specificity. Data are presented for the whole sample and stratified by postmenstrual age and study site.

Performance was comparable across post-menstrual age, sites, and maximum seizure durations (Table 2, Figure 4).

## Discussion

In this real-world and multi-center validation cohort, Clarity demonstrated high performance in automatically detecting and ruling out electroencephalographic seizures in preterm neonates, term neonates, and infants. Therefore, the algorithm may address the critical need for automated, rapid, accurate and round-the-clock seizure detection in these vulnerable patients ^37,46,47^. Clarity was correct 99-100% of the time when ruling out seizures in a recording and identified 89-94% of recordings containing seizures. This indicates that the algorithm may help avoid unnecessary empirical treatment in patients with no seizures and facilitate prompt treatment when a seizure is detected.

Neonatal seizures are difficult to diagnose by clinical observation since they are usually subclinical. Furthermore, it is challenging to distinguish clinical seizures from other non-seizure movements^20,22,48^. This study indicates that Clarity can reliably detect and rule out electroencephalographic seizures in these patients. The algorithm had excellent performance in determining whether 10-second EEG segments contained a seizure, with an AUC of 0.96. It detected 75-83% of individual seizure episodes, depending on seizure duration. For clinical application, performance at the recording level is particularly important since this underlies the algorithm’s ability to inform clinical decisions for individual patients. When ruling out seizures in a recording, Clarity was correct in 99-100% of recordings (negative predictive value), and it correctly detected a seizure in 89-94% of recordings containing at least one seizure (sensitivity). Clarity also had a low rate of false positives per hour of 0.03-0.11 depending on seizure duration. This false positive rate is substantially lower than previous algorithms. For example, a previous study evaluating a neonate seizure detection model ^34^ reported 0.36 and 0.46 false positives per hour in two validation datasets. This low rate of false positives may help avoid “alert fatigue” when Clarity is used in clinical workflows ^49^. It is worth noting that specificity was further increased when not considering as false positives cases in which a seizure was present, but its duration was overestimated by the algorithm, since these might still be clinically relevant. Taken together, these results indicate that across varying thresholds of seizure duration, Clarity consistently maintains high sensitivity and confidence in ruling out seizures while keeping false positives low.

Clarity can reliably detect and rule out seizures in term neonates and infants as well as pre-term neonates, which is an important advance over previous systems. Preterm neonatal EEG patterns differ from those of full-term infants, including in characteristics such as intermittent background activity and rapid maturational changes ^40,50^. Consequently, seizure detection algorithms trained exclusively on EEG data from term neonates may perform worse when applied to preterm data ^51^.

Clarity also performed consistently well on the data from three distinct hospitals which differed in gestational age, recording length, seizure prevalence, and seizure duration, reflecting the variability of demographics, clinical characteristics,and EEG monitoring practices across institutions. Clarity performed well despite this variability, thereby demonstrating generalizability required for clinical adoption.

Continuous conventional EEG monitoring with expert interpretation is the current gold standard for seizure diagnosis, but is often unavailable, especially outside the United States ^52^. Delayed seizure management in the first year of life can have profound consequences given that these seizures are frequently a manifestation of underlying brain injury and are associated with mortality and adverse neurodevelopmental outcomes, including cerebral palsy, epilepsy, and intellectual disability ^7,8,13,53,54^. Clarity’s ability to detect and rule out seizures in preterm neonates, term neonates, and infants may bridge this gap in care, increasing access to high-quality, real-time EEG interpretation. Clarity may enable a more timely bedside diagnosis of seizures in this population, thereby enabling more rapid administration of antiseizure medications. It may also avoid administering unnecessary anti-seizure medications with potential adverse effects in patients who are not experiencing seizures. These improvements in management may ultimately result in better patient outcomes.

Despite its significant advancements, our work has some limitations. First, while the model demonstrated high performance in datasets from three different hospitals, it may still underperform when deployed in institutions with other patient characteristics, comorbidities, or data acquisition protocols. Periodic updates to the algorithm using data from additional institutions may improve its generalizability. Second, since this study was retrospective, we could not test how Clarity would influence clinical practice in real-world settings. Future studies in which the algorithm is used prospectively as part of clinical workflows will be able to determine whether it enables faster seizure recognition and treatment, avoids unnecessary anti-seizure medication administration in neonates without seizures, and if these improvements ultimately yield better long-term neurodevelopmental outcomes. There is data that even with conventional EEG monitoring, seizure management is imperfect with delays to anti-seizure medication administration ^26,55^. Quality improvement initiatives have been shown to improve seizure management, and focusing on both quicker and more efficient seizure detection with devices such as Clarity, as well as linked clinical management approaches, may optimize care ^56^.

Overall, Clarity demonstrated high performance in detecting and ruling out seizures from EEG data in preterm neonates, term neonates, and infants across three hospitals. These results show the value of Clarity for continuous EEG monitoring and seizure management in the first year of life.

## Acknowledgements

We thank Adriane Mueller, Melanie Maki, David Reiber, Chaquitah Jackson and Byron Ramirez-Hamouz for their role in the data transfer.

## Author contributions

CRediT statement: AG: Conceptualization, Data curation, Formal analysis, Investigation, Methodology, Project administration, Resources, Software, Supervision, Validation, Writing - Review & Editing, Writing - Original Draft, Visualization; LT: Conceptualization, Data curation, Formal analysis, Investigation, Methodology, Project administration, Supervision, Visualization, Writing - Original Draft, Writing - Review & Editing, Validation; TP: Writing - Original Draft, Writing - Review & Editing, Conceptualization, Data curation, Formal analysis, Investigation, Methodology, Validation, Visualization, Software; SA: Writing - Original Draft, Writing - Review & Editing, Visualization, Validation, Software, Methodology, Investigation, Formal analysis, Data curation, Conceptualization; CJ: Writing - Review & Editing, Supervision, Investigation, Resources; MV: Writing - Review & Editing, Investigation, Resources, Supervision; RS: Writing - Review & Editing, Investigation, Resources, Supervision; AS: Writing - Review & Editing, Investigation, Resources, Supervision; JS: Writing - Review & Editing, Conceptualization, Supervision; NA: Writing - Review & Editing, Conceptualization, Supervision; CH: Writing - Review & Editing, Conceptualization, Supervision; BK: Writing - Review & Editing, Conceptualization, Supervision, Investigation, Methodology, Funding acquisition, Project administration, Resources, Software, Validation, Writing - Original Draft.

## Source of support

Funding was provided by Ceribell, Inc.

## Conflict of interest

AG, LT, TP, SA and BK are employed by Ceribell (Sunnyvale, California).

NA has received consulting income from Ceribell and UCB Pharma, as well as royalties from Demos Publishing. His institution has received research funding from NINDS and UCB Pharma. CH has received consulting income from Ceribell, Holberg EEG, Takeda Pharmaceuticals and Pendopharm, as well as royalties from Cambridge University Press. His institution has received research funding from NINDS, CIHR, UCB Pharma, Longboard Pharmaceuticals and Takeda Pharmaceuticals.

AS has received consulting income from Ceribell.

## Ethical approval and informed consent

This research study was reviewed and approved by the WCG Institutional Review Board (IRB). The IRB granted a waiver of informed consent for this study.

## Data availability statement

The EEG datasets are not publicly available, and we do not have permission to share the raw data under the terms of the clinical trial agreement and IRB determination.

## Supplementary Material

**Supplementary Table 1:** Qualifications of raters used to obtain seizure labels.

| **Rater** | **Title** | **Fellowship Training** | **Years of Experience** |
| --- | --- | --- | --- |
| Rater 1 | MD, PhD | Epilepsy & Clinical Neurophysiology | 6 |
| Rater 2 | MD | Epilepsy & Clinical Neurophysiology | 18 |
| Rater 3 | MD | Clinical Neurophysiology | 30 |
| Rater 4 | MD | Clinical Neurophysiology | 8 |
| Rater 5 | MD | Clinical Neurophysiology | 10 |
| Rater 6 | MD | Clinical Neurophysiology | 9 |
| Rater 7 | MD | Epilepsy | 12 |

**Supplementary Table 2:**
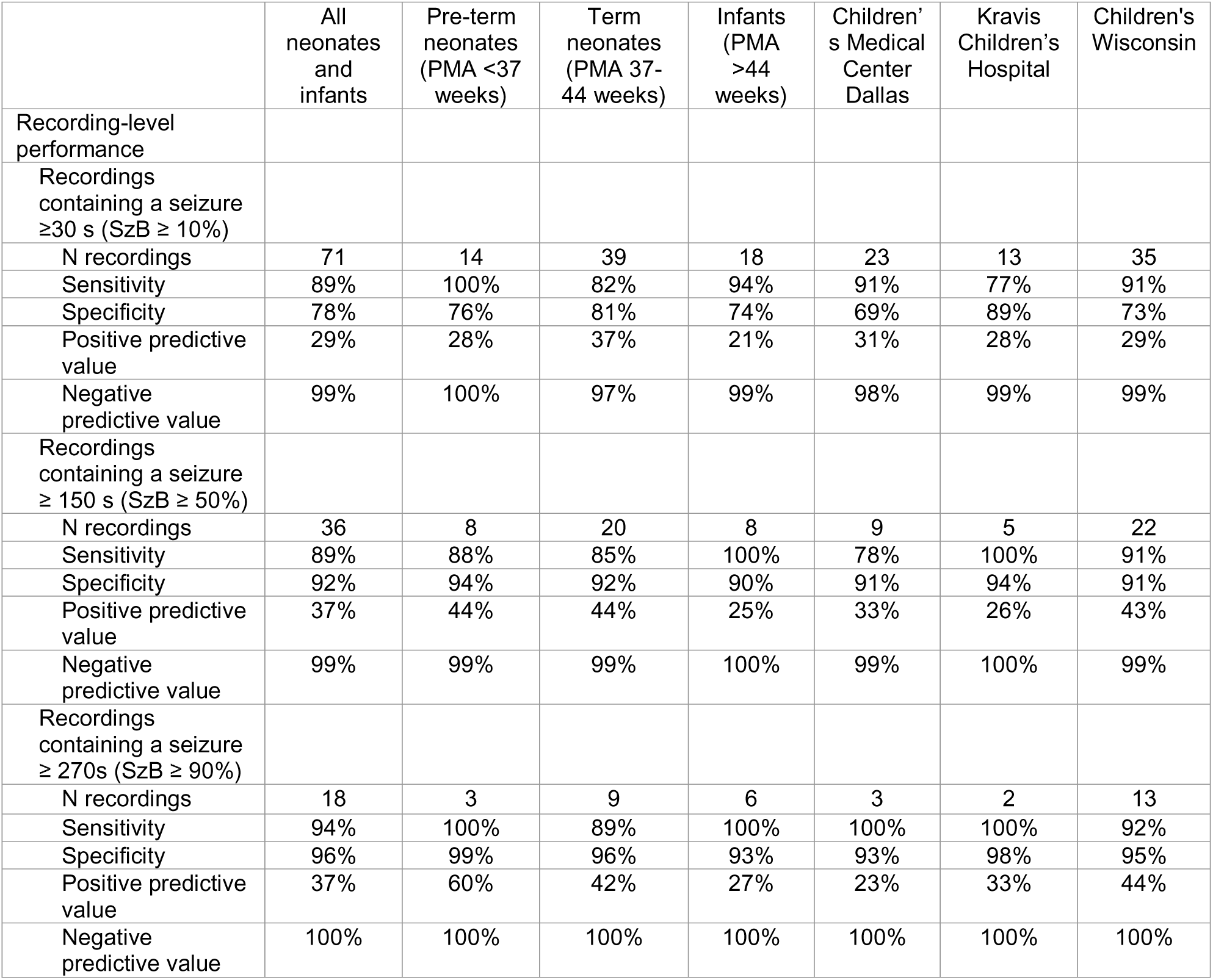
Algorithm performance when excluding recordings that contained seizures with shorter maximum seizure duration than the predefined threshold. We calculated algorithm performance at the recording level after excluding recordings that contained seizures with shorter maximum seizure duration than the predefined threshold. This was done to account for cases where the algorithm correctly detected a seizure but overestimated its duration. An alert in this case might still be clinically relevant and may not be considered a false positive. Here, for instance, given a 270s threshold, a recording with 150s maximum seizure duration with an algorithmically predicted maximum seizure duration of 270s was not counted as a false positive but instead removed from the calculation of performance metrics. Abbreviations: PMA=post-menstrual age, s=seconds, SzB=seizure burden.

